# Plasma Alzheimer’s assays agree on level but much less on individual rates of change

**DOI:** 10.64898/2026.11.16.26368193

**Authors:** Yi Yang, Vanessa Raymont, James B Rowe, Kellen K Petersen, Ahmet Begde, Rachel Brohier, Rosanna McEwen-Smith, Ivan Koychev, the Alzheimer’s Disease Neuroimaging Initiative

## Abstract

**INTRODUCTION:** Serial plasma testing is proposed for monitoring Alzheimer’s disease and for trials, but its reliability for a person’s rate of change is unknown.

**METHODS:** In 393 ADNI participants with three plasma samples over a median 5.0 years, 15 measures of five biomarkers were assayed on aliquots of the same samples, separating shared change from assay-specific variation at each sampling occasion. Results were tested in 1,093 A4/LEARN participants.

**RESULTS:** Assays agreed well on cross-sectional levels (intraclass correlation 0.75–0.95 for pTau217, pTau181, GFAP, NfL) but less on individual rates of change (0.44, 95% CI 0.37–0.50, for pTau217). Assay-specific slope variance was approximately zero, so disagreement was predominantly random. Three-sample single-assay slope reliability was at most 0.40 (0.34–0.46) for pTau217, 0.42 (0.33– 0.51) for pTau181, 0.47 (0.37–0.60) for GFAP, 0.52 (0.40–0.64) for NfL, and 0.10 (0.03–0.20) for Aβ42/40. Two pTau217 measurements had to differ by 65%–131%, depending on the assay, to exceed measurement noise. Detecting 25% slowing of pTau217 change required 924 (598–1,468) to 4,030 (2,091–9,256) participants per arm, or 532 (357–778) to 965 (655–1,431) with averaged measures. In A4/LEARN, single-assay pTau217 reliability was 0.54 (0.46–0.61), and a 7.3-rather than 4.8-year span raised it by 0.10 (0.03–0.22); a fourth sample within it added little.

**DISCUSSION:** With three single-assay samples, much of an individual’s measured change is noise. Longer spans, endpoint replication, and assay choice, rather than additional intermediate samples, are needed for reliable serial testing and trials.

## Introduction

Plasma assays for Alzheimer’s disease pathology, particularly phosphorylated tau 217 (pTau217), have rapidly moved from research into clinical use [1,2]. Serial measurement is now proposed to monitor disease progression, assess response to anti-amyloid treatment, and serve as a trial outcome [3–5]. These uses depend on assumptions for a rarely established quantity: how much of the change observed in an individual’s plasma series is due to disease progression rather than assay variance. Population trajectories are well described, including associations of pTau217 change with amyloid accumulation and subsequent cognitive decline [6,7]. Whether an individual’s rate of change can be trusted is a different issue, determined by measurement noise relative to the biological change accumulating between samples. This matters both clinically, where an apparent rise may fall within measurement noise, and in trials, where noise directly affects the sample size required to detect treatment effects.

The available assays have been compared head-to-head on the same samples and agree closely about a person’s level at a single timepoint, with correlations of 0.80–0.96 between the assays used here for pTau217, %pTau217, pTau181, GFAP, and NfL [8], and of 0.79–0.97 across 13 pTau217 assays and one pTau217 ratio [9]. This agreement about cross-sectional level, however, does not carry over to rates of change. A level is a single measurement compared against the spread of a population; for these biomarkers, that spread is large relative to the noise in one measurement. A rate of change is a difference between measurements divided by an interval. Differencing removes the between-person spread but leaves the measurement noise intact, so agreement about change is poorer than agreement about level, the more so when little change accumulates between samples. Assays can therefore agree well about who has a high level yet disagree about who is changing, and a population trajectory can be estimated well even when few individuals’ rates are distinguishable from zero.

Three properties of measurement are at issue, and they are ordered rather than parallel. Reliability asks how much of an individual’s apparent change is due to a signal of interest; reproducibility, whether a separate measurement of the same blood places that person similarly; and validity, whether the reliable marker change tracks changes in the disease state. The order matters because reliability limits validity: noise weakens the observed correlation with measures of disease progression, such as amyloid PET, CSF biomarkers, MRI, or cognition, even when the underlying changes are closely related.

Under classical test theory, the correlation between an observed rate of change and any reference measure cannot exceed the square root of the rate’s reliability [10–13]. A rate that is 40% reliable can correlate at most 0.63 with a perfectly measured reference, and measurement error in the reference lowers this ceiling further. However, validity has received more attention than the reliability and reproducibility on which it depends; all three are examined here.

True change and measurement noise cannot be separated from a series measured on one assay alone. Measuring the same blood on multiple assays can distinguish shared variation, including true change, from assay-specific error, following generalisability theory [14] and multitrait-multimethod designs [15]. However, pre-assay variation (e.g. in biofluid acquisition, processing or storage) is also shared and therefore not fully distinguishable from true change, making reliability estimates upper bounds. Short-interval repeat samples help resolve this: when little true change is expected, their within-person variation, 7% to 17% for pTau217, pTau181, GFAP, and NfL over days to weeks [16,17], provides a direct benchmark for how much unexplained long-term variation reflects noise.

Four questions are therefore addressed. How reliably do one or multiple assays measure an individual’s rate of change? Is disagreement between assays systematic and correctable by calibration, or random and reducible by further sampling? How large must a change be to exceed measurement noise in each assay’s units? Does reliable change track disease progression across four reference measures? Implications for serial sampling and trials follow.

## Methods

### Participants and samples

Data used in the preparation of this article were obtained from the Alzheimer’s Disease Neuroimaging Initiative (ADNI) database (adni.loni.usc.edu). The ADNI was launched in 2003 as a public-private partnership, led by Principal Investigator Michael W. Weiner, MD. The original goal of ADNI was to test whether serial magnetic resonance imaging (MRI), positron emission tomography (PET), other biological markers, and clinical and neuropsychological assessment can be combined to measure the progression of mild cognitive impairment (MCI) and early Alzheimer’s disease (AD). The current goals include validating biomarkers for clinical trials, improving the generalizability of ADNI data by increasing diversity in the participant cohort, and to provide data concerning the diagnosis and progression of Alzheimer’s disease to the scientific community. For up-to-date information, see adni.loni.usc.edu. Plasma measurements came from the Foundation for the National Institutes of Health Biomarkers Consortium project, which assayed 1,231 stored ADNI samples from 406 participants on multiple commercial and research platforms; a sample identifier shared across platforms for each sampling date confirmed that platforms measured aliquots of the same blood sample.

The analysis included 393 participants with three samples measured on C2N, Fujirebio, ALZpath, Janssen, and Roche. Of these, 342 also had Quanterix measurements, partly from different blood draws. Samples measured only on Quanterix were excluded: all samples of 13 participants (37 samples), and 15 further samples from included participants. The median interval between consecutive samples was 2.5 years (interquartile range, 2.0–3.3), and the median time from first to last sample was 5.0 years (4.0–6.5). Seven participants missing one platform measurement were retained, and samples within seven days were treated as one visit. Diagnosis came from the ADNI merged-dataset visit closest to the first plasma sample, and sex, education, and APOE genotype from the ADNI baseline record.

A second cohort, A4 and its observational companion LEARN, was used to replicate the reliability estimates and to test the effect of sampling design. pTau217 was measured by the Lilly Meso Scale Discovery immunoassay [18] in up to four samples from 1,093 cognitively unimpaired participants: 569 A4 placebo participants (amyloid elevated at screening) and 524 LEARN participants (amyloid negative at screening; 39 amyloid positive in these analyses). Amyloid PET, tau PET, cognition [19], and cerebrospinal fluid (CSF) in a subset were available; florbetapir PET standardised uptake value ratios were converted to Centiloids [20]. Participants gave one (n=91), two (n=315), three (n=610), or four (n=77) samples; the 687 with three or more spanned a median 5.0 years, and the 77 with four a median 7.3 years, the fourth taken during the open-label extension. The solanezumab arm (n=550) formed a sensitivity analysis. A4 repeat samples at week 12 from 1,020 participants (median interval, 85 days) were used to estimate measurement variation over an interval with little expected true change.

### Plasma biomarker measures

Fifteen measures of five biomarkers were analysed. pTau217 was measured by C2N mass spectrometry as concentration and as the percentage of phosphorylated to non-phosphorylated tau 217 (%pTau217), and by three immunoassays: ALZpath (Quanterix Simoa), Fujirebio Lumipulse, and Janssen Simoa. Because %pTau217 divides pTau217 by non-phosphorylated tau 217 from the same sample, pre-analytical variation affecting both should cancel; comparing the two C2N measures tested whether this normalisation improves the measurement of change. pTau181, glial fibrillary acidic protein (GFAP), and neurofilament light chain (NfL) were measured by Roche Elecsys and Quanterix Simoa, and amyloid-β 42/40 (Aβ42/40) by C2N, Fujirebio, Roche, and Quanterix. ALZpath, Janssen, and Quanterix ran samples in duplicate and the others in singlicate, in 12–40 batches per platform [8]. Values below the lower limit of quantification were kept at the value substituted by the laboratory and flagged: 36% of C2N pTau217 observations (94 participants always at 0.65 pg/mL), 3.6% of Fujirebio pTau217 observations, and ≤1% of other measures.

Each measure was log-transformed and standardised (mean 0, SD 1), with Aβ42/40 reversed so that higher values indicated greater abnormality; this removes constant calibration differences between platforms.

### Reference measures

Validity was assessed against four reference measures, chosen at increasing distance from the plasma measurement: CSF pTau181/Aβ42 (Roche Elecsys, University of Pennsylvania) [21]; amyloid PET in Centiloids [22] (University of California Berkeley processing); hippocampal volume on MRI [23], adjusted for intracranial volume (ICV) as hippocampus − 0.0017 × (ICV − 1,498,858 mm³); and cognition. Cognition was measured by the Alzheimer’s Disease Assessment Scale cognitive subscale, 13-item version (ADAS-Cog13) [24,25], the modified Preclinical Alzheimer Cognitive Composite (mPACC; digit-symbol version) [26], and the Mini-Mental State Examination (MMSE) [27]. Change in each was the least-squares slope of assessments within the plasma sampling period ±180 days, requiring at least two. Amyloid status was defined at the first plasma sample (> 20 Centiloids [28] on the nearest scan within 180 days). CSF served only as a reference for ranking plasma rates of change; within-person associations between plasma and CSF are reported separately.

### Statistical analysis

The analysis assessed agreement between platforms about level and change, divided disagreement about change into systematic and occasion-level components, and derived from these the reliability of an individual slope, the minimal detectable change, and trial sample sizes. A4 analyses tested the underlying assumptions, sensitivity analyses tested analytical choices and model form, and validity was assessed against the reference measures. Analyses used R 4.5.1 [29] with lme4 [30] and lcmm [31], with the same code for both cohorts; full specifications are in the Supplementary Methods.

### Agreement between platforms

Agreement about level was assessed across all samples measured on both platforms, on the log scale, by Pearson correlation, mean difference with 95% limits of agreement [32], and intraclass correlation (ICC) after removing platform means, and by the generalisability coefficient [14] of one measurement on one platform from a crossed random-effects model (participant, platform, participant by platform, sample, platform by visit, and batch). Agreement about change was assessed between per-platform least-squares slopes, the quantity whose reliability is estimated, by Pearson and Spearman correlations, ICCs [33], and Cohen’s kappa for slope tertiles [34].

### Decomposition of change and reliability of an individual slope

A joint linear mixed model [35] was fitted for each biomarker, with fixed effects for platform, time, and their interaction, and random effects for a participant intercept and slope shared across platforms (true level and change), a participant-by-platform intercept and slope (platform-specific deviations), and batch. A likelihood-ratio test with boundary correction [36,37] tested the platform-specific slope variance. For pTau217, power to detect this variance was estimated by simulation from the reduced model’s components on the observed design, adding platform-specific slope SDs of up to 0.8 times the true between-person slope SD and refitting both models. Because this variance was approximately zero for every biomarker, the reduced model without it was used for the bootstrap and the stratified and sensitivity analyses, and the full and reduced models gave the same validity correlations to three decimal places. Change was modelled as linear because three samples cannot separate individual curvature from measurement variation; the A4 four-sample analysis tested this for the mean trajectory.

Slope reliability, the proportion of observed slope variance due to true between-person differences in change, was estimated as var(true slope) / [var(true slope) + var(platform-specific slope) + var(residual)/SST], where SST (spread of sampling times) is the sum of squared deviations of a participant’s sampling times from their mean [12,13]; the median participant SST of 12.9 years² was used. Because variation shared by all platforms at a blood draw cannot be separated from true change in the joint model, this model gives upper bounds. Adding a random effect for each plasma sample divides occasion-level variance into shared and assay-specific parts and gives lower bounds, identifiable only for pTau217, which has five platforms. Fitting each measure alone, treating all variation around a participant’s line as error, gives the like-for-like comparison with the second cohort. For the mean of k platforms, the error variance, or for the lower bound its assay-specific part, was divided by k. Reliability was also estimated by baseline amyloid status.

Percentile 95% confidence intervals came from 1,000 participant-level bootstrap resamples with model refitting [38], with differences computed within resamples. Residuals were inspected against fitted values and time. Paired comparisons are descriptive and were not adjusted for multiple testing.

### Short-interval test-retest and direct tests of sampling design

Three analyses in A4 tested the assumptions that the change-model residual represents measurement variation and that reliability depends on sampling design only through SST. First, baseline and week-12 samples formed a test-retest pair, summarised by the within-person coefficient of variation (CV) [39], ICC(2,1) (absolute agreement) [33,40], and Bland-Altman limits [32]. In participants with a fourth sample, the test-retest within-person variance was compared with the residual around a line through the baseline, week-240, and extension samples, and curvature of the mean trajectory was tested with a quadratic term. Second, each participant’s slope was recalculated from each pair of their three samples (SST 0.03, 10.24, and 11.32 against 14.40 years² for all three), and its correlation with concurrent amyloid PET change (Spearman, with Pearson as a check) was compared with that predicted from reliability; the same relation was examined across bands of observed SST in both cohorts. Third, in participants with a fourth sample, four designs (samples 1–3; 1, 2, and 4; 1, 3, and 4; all four) separated the effects of a longer span, an added sample, and a moved intermediate sample. Because both arms received open-label solanezumab before the fourth sample, the placebo arm was primary for the design comparisons, so that its first three samples were untreated; the four-sample residual test used both arms.

### Sensitivity and alternative-model analyses

Sensitivity analyses repeated the pTau217 decomposition under five treatments of C2N values at the lower limit of quantification (as reported; excluding the 94 participants always at the limit; excluding values at the limit; substituting half the limit; and multiple imputation from the other platforms [41]). The %pTau217 comparison was repeated without values at the limit and in participants never at the limit. For every biomarker, analyses were repeated adjusting for age, sex, APOE ε4 carriage, and diagnosis at the first sample, each interacting with time, and under two alternative scalings (untransformed values, and log values standardised on the first visit). All samples were assayed in January–March 2024, so storage time was set by collection date; first-sample levels were regressed on storage time, adjusted for age, amyloid status, and diagnosis, to estimate any degradation bias in the rate of change, and each participant’s slope was regressed on storage time, which cannot be separated from enrolment period or assay batch. Model form was tested with a latent-process mixed model [31], treating all platforms as noisy measures of one process, and a multitrait-multimethod model [15] of the four analytes measured by Roche and Quanterix, testing whether disagreement between vendors was shared across analytes.

### Minimal detectable change and its consequences

Measurement noise was the within-person residual SD from a mixed model with participant intercepts and slopes, fitted to each measure on the standardised log scale. The 95% minimal detectable change (MDC95) [42] was 1.96 × √2 × this SD between two measurements, and 1.96 × the slope standard error (SD/√SST) for a slope at the median SST, converted to fold change on the original scale. From these, the years needed for the mean amyloid-positive rate of change to exceed MDC95, and the proportions of all participants whose own slope exceeded the slope MDC95 in either direction, were calculated. The analytical share of within-person variance was the squared ratio of the quality-control CV [8] to the within-person CV; the remainder comprised pre-analytical variation, short-term biological variation, and departure from linearity.

Reliability was calculated for schedules differing in sample number, spacing, and span, including the span needed for reliabilities of 0.70, 0.80, and 0.90 with two, three, four, or six evenly spaced samples. Participants per arm were calculated for a two-arm trial detecting 25% or 50% slowing of the mean amyloid-positive rate (80% power, two-sided α=0.05), from individual slope variance under the trial schedule [43] with amyloid-positive variance components, for one platform and for the mean of platforms. Change from baseline to the final sample, with whole-cohort variance components, was used as an alternative outcome. These calculations show how assay and design change requirements in an ADNI-like population rather than prescribe a trial.

### Validity

The pooled rate of change (the shared participant slope from the multi-platform model) and each single-platform slope were correlated with concurrent change in each reference measure, using Spearman correlation because several slope distributions were heavy-tailed and no extreme values were removed; signs were set so that positive correlations indicated worsening. For pTau217 and Aβ42/40, the reduced model was refitted with each measure omitted in turn, and the resulting pooled slope was compared with the all-platform pooled slope and with concurrent amyloid PET change.

## Results

### Participants

Among 393 participants, 146 (37%) were amyloid positive at baseline; of 247 negative participants, 51 (21%) became positive between their first and last plasma samples, and 14 (6%) at a later scan. Amyloid-positive participants were older, more often APOE ε4 carriers (53% vs 22%) and cognitively impaired, and had shorter sampling spans (median 4.2 vs 5.5 years; Table 1). C2N pTau217 was at its quantification limit in 54% of amyloid-negative and 5% of amyloid-positive samples, and 90 amyloid-negative and 4 amyloid-positive participants were at the limit at every sample (Table 1). Concurrent slopes were available for all participants for amyloid PET and cognition, 371 (94%) for hippocampal volume, and 207 (53%) for CSF. Assay characteristics are in Supplementary Table S1; Figure 1 shows the design and pTau217 series, with other biomarkers in Supplementary Figure S1.

**Figure 1.**
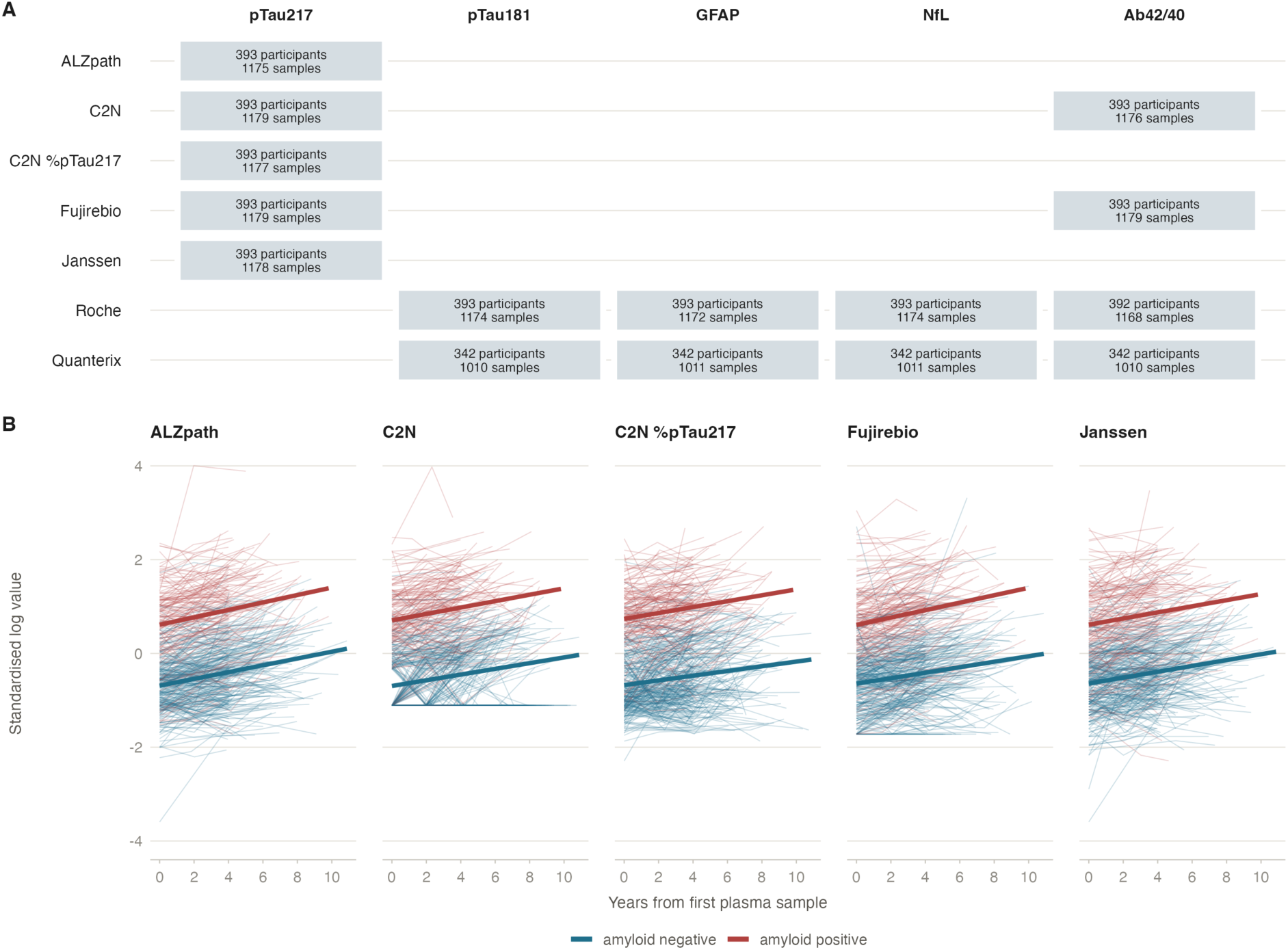
Study design and individual pTau217 trajectories. (A) Platforms and biomarkers: each cell gives the number of participants and plasma samples measured on that platform for that biomarker. All 393 participants provided three samples over a median of 5.0 years, and every platform assayed an aliquot of the same blood sample. C2N reports pTau217 both as a concentration and as the percentage of phosphorylated to non-phosphorylated tau (%pTau217). (B) Every participant’s pTau217 series on each platform. Values are log-transformed and standardised within measure so that higher is more abnormal; thin lines are individual participants, heavy lines the ordinary least-squares trend by amyloid PET status at the first sample (Centiloid > 20). The vertical axis is truncated at ±4 standardised units.

**Table 1.**
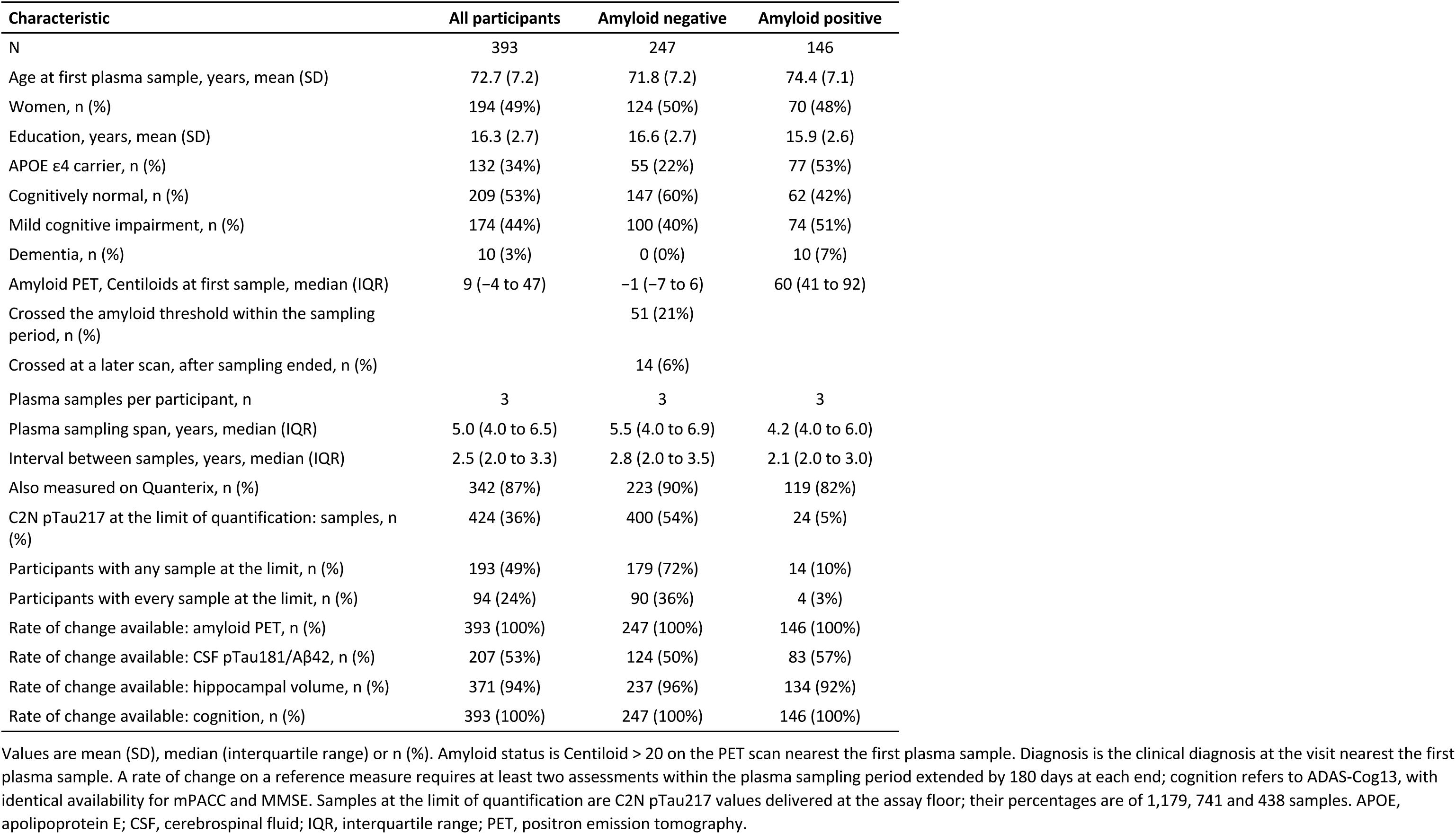
Characteristics of the 393 participants, overall and by amyloid PET status at the first plasma sample.

### Platforms agree about a participant’s level

Platforms measuring the same biomarker on the same aliquot generally agreed on a participant’s relative level after fixed calibration differences were removed. Centred intraclass correlations were 0.95 for NfL and 0.94 for GFAP (Roche vs Quanterix), 0.78 for pTau181, and 0.75–0.92 across the five pTau217 measures. The closest pTau217 pairs were C2N vs %pTau217 (0.925), two readouts of the same assay, and ALZpath vs Janssen (0.922), two separate assays; %pTau217 was least concordant with the other measures (0.75–0.77). Aβ42/40 was the exception, with correlations of only 0.28–0.51 across its six platform pairs (Supplementary Table S2, Figure S2).

Agreement in relative level did not mean that platforms reported similar absolute values. Fixed calibration offsets were large: C2N reported pTau217 concentrations 32.32-fold those of Janssen, and Quanterix pTau181 17.90-fold those of Roche. More relevant to agreement was the variation around these offsets. The SD of log differences between pTau217 platforms ranged from 0.27 to 0.60, corresponding after removal of the mean offset to 95% limits of agreement of 0.59–1.69-fold and 0.31–3.23-fold; for NfL it was 0.15, or 0.75–1.34-fold.

Variance decomposition explained this pattern (Figure 2A, Table 2). True between-participant differences in level accounted for 76% of pTau217 variance, 77% of GFAP, 72% of NfL, and 66% of pTau181, but only 30% of Aβ42/40 variance; for Aβ42/40, participant-by-platform (27%) and residual variation (29%) were each as large as the signal. Accordingly, generalisability coefficients for one measurement on one platform were 0.79 (95% CI 0.77–0.82) for pTau217, 0.81 (0.78–0.85) for GFAP, 0.76 (0.71–0.80) for NfL, 0.70 (0.67–0.75) for pTau181, and 0.32 (0.26–0.40) for Aβ42/40 (Supplementary Tables S3 and S4). Analytical batch accounted for ≤2% of variance except for Aβ42/40 (7%). The sample effect, variation shared across platforms measuring the same sample and therefore attributable to the participant that day or pre-analytical handling rather than any assay, was largest for NfL (19%) and GFAP (13%).

**Figure 2.**
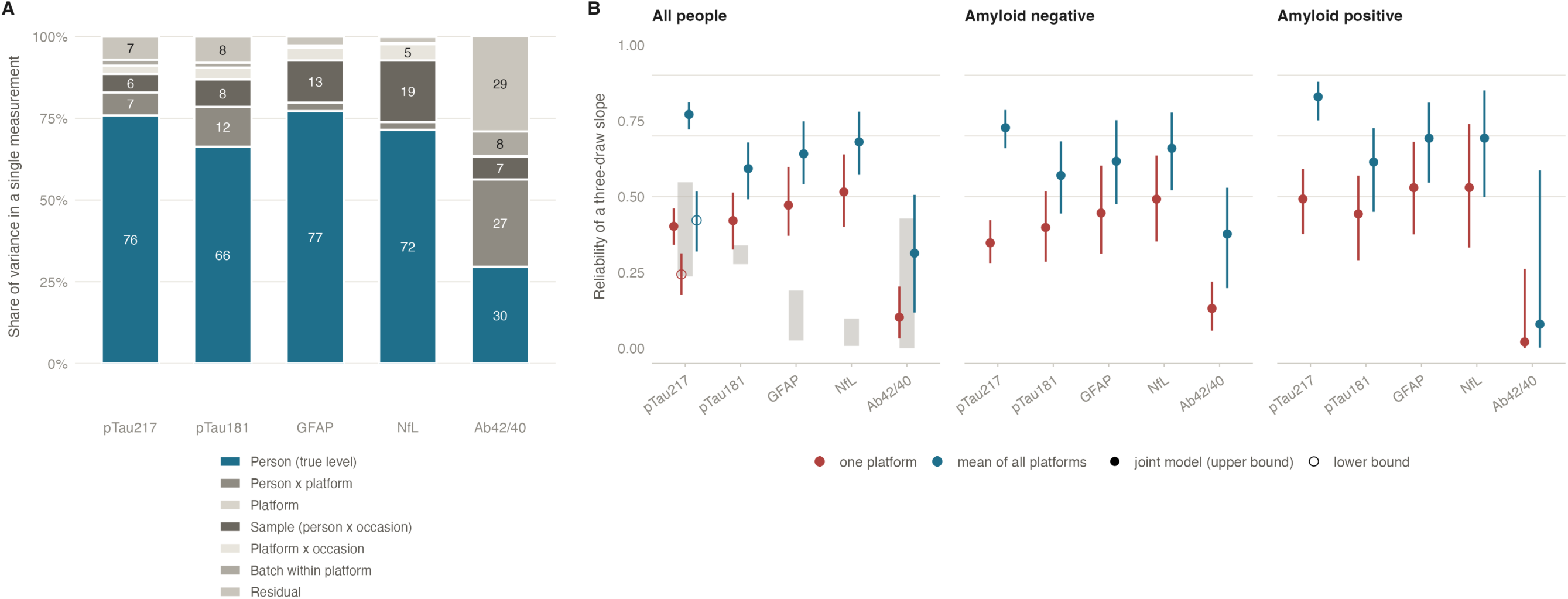
Platforms agree about level but not about change. (A) Sources of variance in a single measurement, from a crossed random-effects model per biomarker: the participant component (blue) is between-participant variance in true level; every other component is error for one measurement on one platform (sample is the participant × visit effect shared by all platforms assaying that aliquot). Segments under 5% are unlabelled. (B) Reliability of an individual’s three-sample rate of change, defined as the share of the variance of an observed slope that is true between-participant variance in rate of change, for a slope from one platform and for the mean of all platforms measuring that biomarker, in all participants and by amyloid PET status at the first sample. Vertical lines are 95% percentile intervals from 1,000 bootstrap resamples of participants. The platform-specific slope variance was estimated at zero for every biomarker, so the gain from averaging platforms comes from averaging assay-specific measurement noise. Filled symbols are the joint-model values, which treat variation shared across platforms at a blood draw as true change and are upper bounds; open symbols (pTau217, all participants) are the lower bounds from the model with a participant-by-sample effect; grey bands span the reliability of each measure fitted alone, from the lowest to the highest platform.

**Table 2.** Sources of variance in a single measurement and reliability of an individual rate of change, per biomarker.

|  | **pTau217** | **pTau181** | **GFAP** | **NfL** | **Aβ42/40** |
|---|---|---|---|---|---|
| Platforms, n | 5 | 2 | 2 | 2 | 4 |
| Level: participant (true level) | 76 | 66 | 77 | 72 | 30 |
| Level: participant × platform | 7 | 12 | 3 | 2 | 27 |
| Level: sample (shared by platforms) | 6 | 8 | 13 | 19 | 7 |
| Level: batch within platform | 2 | 1 | 1 | 0 | 7 |
| Level: residual | 7 | 8 | 3 | 2 | 29 |
| G coefficient, one measurement | 0.79 (0.77 to 0.82) | 0.70 (0.67 to 0.75) | 0.81 (0.78 to 0.85) | 0.76 (0.71 to 0.80) | 0.32 (0.26 to 0.40) |
| SD of true rate of change, per year | 0.068 | 0.080 | 0.082 | 0.101 | 0.055 |
| SD of platform-specific rate of change | 0.000 | 0.000 | 0.000 | 0.000 | 0.000 |
| Residual SD | 0.30 | 0.34 | 0.31 | 0.35 | 0.58 |
| Slope reliability, one platform (joint model, upper bound) | 0.40 (0.34 to 0.46) | 0.42 (0.33 to 0.51) | 0.47 (0.37 to 0.60) | 0.52 (0.40 to 0.64) | 0.10 (0.03 to 0.20) |
| Slope reliability, one platform (lower bound) | 0.24 (0.18 to 0.31) | not estimable | not estimable | not estimable | not estimable |
| Slope reliability, each measure fitted alone, lowest to highest platform | 0.24 (0.12 to 0.34) to 0.55 (0.33 to 0.67) | 0.28 (0.12 to 0.41) to 0.34 (0.12 to 0.48) | 0.03 (0.00 to 0.19) to 0.19 (0.02 to 0.32) | 0.01 (0.00 to 0.25) to 0.10 (0.00 to 0.38) | 0.00 (0.00 to 0.44) to 0.43 (0.05 to 0.64) |
| Slope reliability, mean of platforms (upper bound) | 0.77 (0.72 to 0.81) | 0.59 (0.49 to 0.68) | 0.64 (0.54 to 0.75) | 0.68 (0.57 to 0.78) | 0.31 (0.12 to 0.51) |
| Slope reliability, mean of platforms (lower bound) | 0.42 (0.32 to 0.52) | not estimable | not estimable | not estimable | not estimable |
| Difference, mean of platforms minus one platform (upper bound) | 0.369 (0.349 to 0.380) | 0.172 (0.164 to 0.172) | 0.169 (0.151 to 0.172) | 0.165 (0.141 to 0.172) | 0.211 (0.086 to 0.302) |
| Slope reliability, one platform, amyloid negative | 0.35 (0.28 to 0.42) | 0.40 (0.29 to 0.52) | 0.45 (0.31 to 0.60) | 0.49 (0.35 to 0.64) | 0.13 (0.06 to 0.22) |
| Slope reliability, one platform, amyloid positive | 0.49 (0.38 to 0.59) | 0.44 (0.29 to 0.57) | 0.53 (0.38 to 0.68) | 0.53 (0.33 to 0.74) | 0.02 (0.00 to 0.26) |
| Slope ICC, one platform / mean of platforms | 0.44 (0.37 to 0.50) / 0.80 (0.75 to 0.84) | 0.59 (0.48 to 0.68) / 0.74 (0.65 to 0.81) | 0.81 (0.71 to 0.89) / 0.89 (0.83 to 0.94) | 0.88 (0.81 to 0.93) / 0.94 (0.90 to 0.96) | 0.18 (0.10 to 0.28) / 0.47 (0.31 to 0.61) |
| Slope ICC, immunoassays only (ALZpath, Fujirebio, Janssen), one platform / mean of platforms | 0.61 (0.53 to 0.69) / 0.82 (0.77 to 0.87) |  |  |  |  |
| **Second cohort, A4 and LEARN (Lilly MSD, one platform): slope reliability; SD of true rate of change, residual SD** |  |  |  |  |  |
| Slope reliability, A4 placebo (n = 569) | 0.60 (0.50 to 0.68); 0.093, 0.28 |  |  |  |  |
| Slope reliability, LEARN (n = 524) | 0.35 (0.21 to 0.48); 0.050, 0.24 |  |  |  |  |
| Slope reliability, placebo + LEARN (n = 1093) | 0.54 (0.46 to 0.61); 0.078, 0.26 |  |  |  |  |
| Slope reliability, A4 solanezumab (n = 550) | 0.58 (0.45 to 0.68); 0.096, 0.30 |  |  |  |  |

Level components are percentages of the variance of a single standardised log measurement from the crossed random-effects model (the platform component, estimated at zero for every biomarker, and the platform × visit component, 0.3% to 5.0%, are omitted). The G coefficient is the participant variance divided by the sum of the participant, participant × platform, sample and residual variances. Slope quantities are from the multi-platform change model on the standardised log scale; SDs are in standardised units per year. Slope reliability is the share of the variance of an observed three-sample slope that is true between-participant variance in rate of change, for the observed design (median spread of sampling times 12.9 years²), on one platform and for the mean of all platforms measuring that biomarker. The platform-specific rate of change was estimated at zero for every biomarker. Reliabilities from the joint model are upper bounds; the lower bounds come from the model with a participant-by-sample effect and are estimable only for pTau217. Each measure fitted alone estimates its own slope variance and residual, with all variation around a participant's line counted as error, and is the like-for-like comparison with the second cohort (Supplementary Table S9c). The immunoassay-only ICC excludes the two C2N measures. The A4 and LEARN SDs are on that cohort's own standardised scale and are not comparable with the ADNI SDs. G, generalisability coefficient; ICC, intraclass correlation of per-participant slopes across platforms. Intervals are 95% percentile intervals from 1,000 bootstrap resamples of participants. Point estimates are differences between the observed estimates; intervals are percentiles of the same difference computed within each bootstrap resample. Differences are given to three decimal places because two would not separate the interval from the estimate. With two platforms the gain from averaging cannot exceed 3 − 2√2 = 0.172, which is why the intervals for pTau181, GFAP and NfL share that upper limit. The pooled reliability is bounded above and below only for pTau217, the one biomarker measured on enough platforms to identify an occasion effect shared between them; for the others only the upper bound is estimable. All paired differences are in Supplementary Table S26.

### Platforms agree much less about a participant’s change

Agreement about rate of change was substantially lower than agreement about level for every biomarker (Supplementary Table S5, Figure S3). Three-sample slopes correlated between platforms at 0.88 for NfL, 0.81 for GFAP, and 0.61 for pTau181. For pTau217, correlations were 0.53–0.73 among the three immunoassays, 0.38–0.46 between C2N concentration and the immunoassays, and only 0.15–0.26 for %pTau217 against the immunoassays; Aβ42/40 correlations were 0.10–0.26.

Single-platform slope intraclass correlations were 0.44 (95% CI 0.37–0.50) for pTau217, 0.59 (0.48– 0.68) for pTau181, 0.81 (0.71–0.89) for GFAP, 0.88 (0.81–0.93) for NfL, and 0.18 (0.10–0.28) for Aβ42/40, increasing to 0.80 (0.75–0.84), 0.74 (0.65–0.81), 0.89 (0.83–0.94), 0.94 (0.90–0.96), and 0.47 (0.31–0.61) when platforms were averaged. Agreement among pTau217 platforms on slope tertiles was slight to moderate (κ=0.17–0.47). Thus, %pTau217 was the least reproducible pTau217 measure for change as well as level, contrary to the expectation that normalisation by non-phosphorylated tau would reduce pre-analytical variation.

### C2N %pTau217 does not improve measurement of change

Normalising pTau217 to non-phosphorylated tau did not make C2N %pTau217 more reliable than C2N pTau217 for tracking change. The two C2N measures agreed with each other on a participant’s rate of change at 0.76. Across separate comparisons with the three pTau217 immunoassays (ALZpath, Fujirebio, and Janssen), however, correlations were only 0.15–0.26 for C2N %pTau217, compared with 0.38–0.46 for C2N pTau217. C2N %pTau217 also had the loosest agreement about level among the pTau217 measures (intraclass correlation 0.75–0.77).

The poorer performance of C2N %pTau217 was not explained by greater measurement noise. Its within-person coefficient of variation was 30% versus 31% for C2N pTau217 (difference 0.85, 95% CI −1.00 to 2.78), and its minimal detectable change was 126% versus 131% (difference 5.00, 95% CI −5.91 to 16.74). Instead, the measures differed in the size of change relative to similar noise: among amyloid-positive participants, C2N %pTau217 rose by 5.2% per year versus 8.1% for C2N pTau217. At these mean rates, 16 versus 11 years between samples were needed for change to exceed measurement noise (difference 5.28 years, 95% CI 2.67–11.01), and a trial using the C2N %pTau217 slope required 4.36 (95% CI 2.22–9.25) times as many participants as one using the best pTau217 measure. Without samples at the quantification limit, the within-person CV fell from 31% to 26% for C2N pTau217 and from 30% to 25% for %pTau217, the minimal detectable change from 131% to 101% and from 126% to 96%, and the years needed to exceed it from 11 to 9 and from 16 to 14; the two C2N slopes agreed at 0.67 without floor samples and 0.72 in the 200 participants never at the floor (Supplementary Table S6a). On none of these measures did %pTau217 improve the measurement of change in any sample. Thus, normalising to non-phosphorylated tau added no measurable noise to a single measurement and removed none, while C2N %pTau217 showed a mean rate of change 36% smaller.

### The disagreement about change is predominantly random rather than systematic

To distinguish systematic platform disagreement from random measurement variation, the mixed model separated each participant’s shared true rate of change from platform-specific slope deviations, representing stable differences in the rate assigned to that participant. For all five biomarkers, the variance of these deviations was estimated at the boundary of zero (boundary-corrected likelihood-ratio p=0.50 in every case; Supplementary Table S7), providing no evidence that any platform systematically made a participant appear to change faster or slower than another.

To determine whether an important systematic difference could have escaped detection, power was estimated by simulation on the observed design. The test had 80% power once the platform-specific slope SD reached 0.27 of the true between-person slope SD (0.02 standardised units/year), with type I error of 0.05 (Supplementary Table S8, Figure S4). An effect at this threshold would account for only 4.8% of the error in a one-platform slope and reduce reliability from 0.40 to 0.39. Smaller systematic differences cannot be excluded, but none large enough to materially affect an individual slope was detected. Instead, disagreement arose predominantly at individual measurement occasions (Supplementary Table S9). Residual variance was similar across platforms and biomarker ranges, except for larger NfL residuals at high values and larger Aβ42/40 residuals on Fujirebio and C2N than Roche and Quanterix (Supplementary Figure S5).

To test whether this conclusion depended on the mixed-model form, two alternative models were fitted. A latent-process model allowing each platform its own link and error produced harmonised participant slopes correlating with pooled mixed-model slopes at 0.98 for pTau217, 1.00 for GFAP and NfL, 0.96 for pTau181, and 0.75 for Aβ42/40 (Supplementary Table S10). The lower Aβ42/40 agreement reflected precision weighting toward Quanterix, whose latent-scale error (0.45) was 21%–30% of that of the other three platforms (1.5–2.2). Latent-scale error ranked the pTau217 measures as ALZpath (0.39), Janssen (0.43), C2N pTau217 (0.44), Fujirebio (0.48), and C2N %pTau217 (0.61); for pTau181, Quanterix error (0.71) was 1.79 times Roche error (0.40; Supplementary Table S11).

To test for a vendor or matrix effect shared across analytes, a multitrait-multimethod model was fitted to the four analytes measured by both vendors. No participant-by-vendor effect shared across analytes was detected at either level or slope, nor was a per-sample vendor effect detected. Instead, vendor disagreement was estimated as wholly analyte-specific (participant-by-analyte-by-vendor, 15% of level variance), with no detectable common rate of change across the four analytes within a participant (Supplementary Table S12). Thus, Roche–Quanterix disagreement was consistent with analyte-specific calibration rather than a shared matrix or vendor effect, supporting separate treatment of platforms within each biomarker.

### Averaging across platforms improves the reliability of individual change

To quantify how much of an individual’s observed rate of change was reliable, the mixed-model variance decomposition separated true between-person slope variance from measurement error. Reliability was the proportion attributable to true slope variance, with measurement error determined by the residual variance and spread of sampling times (SST). Under the observed design of three samples over a median 5.0 years, single-platform reliability was 0.40 (95% CI 0.34–0.46) for pTau217, 0.42 (0.33–0.51) for pTau181, 0.47 (0.37–0.60) for GFAP, 0.52 (0.40–0.64) for NfL, and 0.10 (0.03–0.20) for Aβ42/40 (Table 2, Figure 2B). These values are upper bounds from the joint model, which counts variation shared by all platforms at a draw as true change; for pTau217, the lower bound from the model with a sample effect was 0.24 (95% CI 0.18–0.31). Each measure fitted alone, with all variation around a participant’s line counted as error, gave 0.24 (0.12–0.34) to 0.55 (0.33– 0.67) across the five pTau217 measures, 0.28 to 0.34 for pTau181, 0.03 to 0.19 for GFAP, 0.01 to 0.10 for NfL and 0.00 to 0.43 for Aβ42/40 (intervals in Table 2); against the joint-model value, C2N pTau217 and C2N %pTau217 were lower by 0.13 and 0.17 with intervals excluding zero, whereas ALZpath, Janssen and Fujirebio showed no detectable difference (−0.05, −0.10 and 0.15, intervals including zero; paired differences in Supplementary Table S9b).

The four proteins showed no detectable difference: the largest paired difference, NfL versus pTau217, was 0.11 (95% CI −0.02 to 0.25), whereas Aβ42/40 was lower than all four (vs NfL, difference 0.41, 0.26–0.56). In the single-measure fits, GFAP and NfL were the lowest of the proteins (Roche 0.19 and 0.10, Quanterix 0.03 and 0.01, with upper interval limits of 0.19 to 0.38), each below its joint-model value by 0.28 to 0.51 with paired intervals excluding zero. True slope SDs were 0.07–0.10 standardised units/year for the proteins and 0.05 for Aβ42/40, compared with occasion-level residual SDs of 0.30–0.35 and 0.58, respectively. Thus, at least 48%–60% of the variation in individual slopes observed on one platform was measurement noise.

To determine how much reliability improves by averaging measurements, slopes were averaged across all platforms measuring each biomarker. Reliability increased to 0.77 (0.72–0.81) for pTau217 across five measures, and to 0.59 (0.49–0.68), 0.64 (0.54–0.75), 0.68 (0.57–0.78), and 0.31 (0.12– 0.51) for pTau181, GFAP, NfL, and Aβ42/40 across their two to four measures. With two platforms the gain cannot exceed 0.17, and the upper limits of the gain intervals for pTau181, GFAP and NfL in Table 2 reach this value. All of these pooled reliabilities are upper bounds. Because every platform measured the same blood sample, averaging removes only the assay-specific part of the occasion-level variation, and the part shared across platforms at a draw remains. For pTau217, the one biomarker with enough platforms to estimate that shared part, it was 30% (95% CI 25–36%) of the occasion-level variance, and the reliability of the mean of platforms with it retained was 0.42 (0.32– 0.52), so the pooled reliability of pTau217 lies between 0.42 and 0.77 (Table 2). Among the three pTau217 immunoassays alone, which exclude the two C2N readouts of one mass-spectrometry measurement, the slope intraclass correlation was 0.61 (0.53–0.69) for one platform and 0.82 (0.77– 0.87) for their mean, against 0.44 and 0.80 across all five measures (paired differences 0.17, 0.12– 0.22, and 0.03, −0.01 to 0.06).

To determine whether reliability differed by amyloid status, estimates were stratified at the first plasma sample. For pTau217, reliability was higher in amyloid-positive than amyloid-negative participants: 0.49 (0.38–0.59) versus 0.35 (0.28–0.42) on one platform and 0.83 (0.75–0.88) versus 0.73 (0.66–0.79) for the mean of platforms. Paired differences were 0.14 (0.01–0.27) and 0.10 (0.01– 0.19), respectively. True slope variance was 34% larger and residual variance 26% smaller in the positive group, so both contributed. For pTau181, GFAP, and NfL, reliabilities were 0.40–0.49 in amyloid-negative and 0.44–0.53 in amyloid-positive participants, with differences of 0.04 (−0.15 to 0.22), 0.08 (−0.14 to 0.30), and 0.04 (−0.20 to 0.29), respectively. For Aβ42/40, reliability was 0.13 (0.06–0.22) in amyloid-negative and 0.02 (0.00–0.26) in amyloid-positive participants, a difference of −0.11 (−0.19 to 0.14); in the positive group, true slope variance was 21% of that of the negative group, consistent with Aβ42/40 having already fallen by PET-detectable amyloid (Supplementary Table S13).

To test robustness to analytical choices, all sensitivity analyses were repeated (Supplementary Tables S6b–S6d). For pTau217, five treatments of C2N values at the quantification limit yielded single-platform reliabilities of 0.37–0.40 and mean-of-platform reliabilities of 0.74–0.77. Cross-platform imputation of log C2N pTau217 from the same sample’s three immunoassays (R²=0.77, residual SD=0.31) did not materially alter the result. Adjustment for age, sex, APOE ε4, and diagnosis reduced between-participant level variance by 19% to 35% for every biomarker but changed slope variances by 3% or less and residual variances by less than 1%, shifting reliability by 0.006 or less.

Standardising on the first rather than all visits gave identical results to two decimals except for Aβ42/40 (0.11 against 0.10 on one platform and 0.33 against 0.31 for the mean of platforms). Standardising untransformed values changed one-platform reliability by 0.03–0.08 for four biomarkers but reduced NfL from 0.52 to 0.21. Storage time, 5.5 to 13.6 years at the first sample, showed no detectable association with the level of any protein biomarker after adjustment for age, amyloid status and diagnosis; the bias it implies in the rate of change lay within −67% to 14% of the mean amyloid-positive rate for ALZpath pTau217 and −73% to 10% for Fujirebio, with point estimates in the direction opposite to degradation for 9 of 11 protein measures (Supplementary Table S14).

### Individual change must be large enough to exceed measurement variation

To quantify measurement variation relative to biological change, within-person coefficients of variation (CVs), comprising all variation in a single measurement other than the participant’s linear trajectory, were expressed on the delivered scale (Table 3, Figure 3). CVs were 18% for ALZpath pTau217, 21% for Janssen, 24% for Fujirebio, 31% for C2N pTau217, and 30% for C2N %pTau217; 15% and 21% for Roche and Quanterix pTau181; 18–21% for GFAP and NfL; and 8% for Aβ42/40 on C2N, Quanterix, and Roche and 16% on Fujirebio. In comparison, amyloid-positive participants changed by 5.2–9.0%/year on pTau217 measures, 3.7–6.7%/year on pTau181, GFAP, and NfL, and −0.7 to 0.2%/year on Aβ42/40.

**Figure 3.**
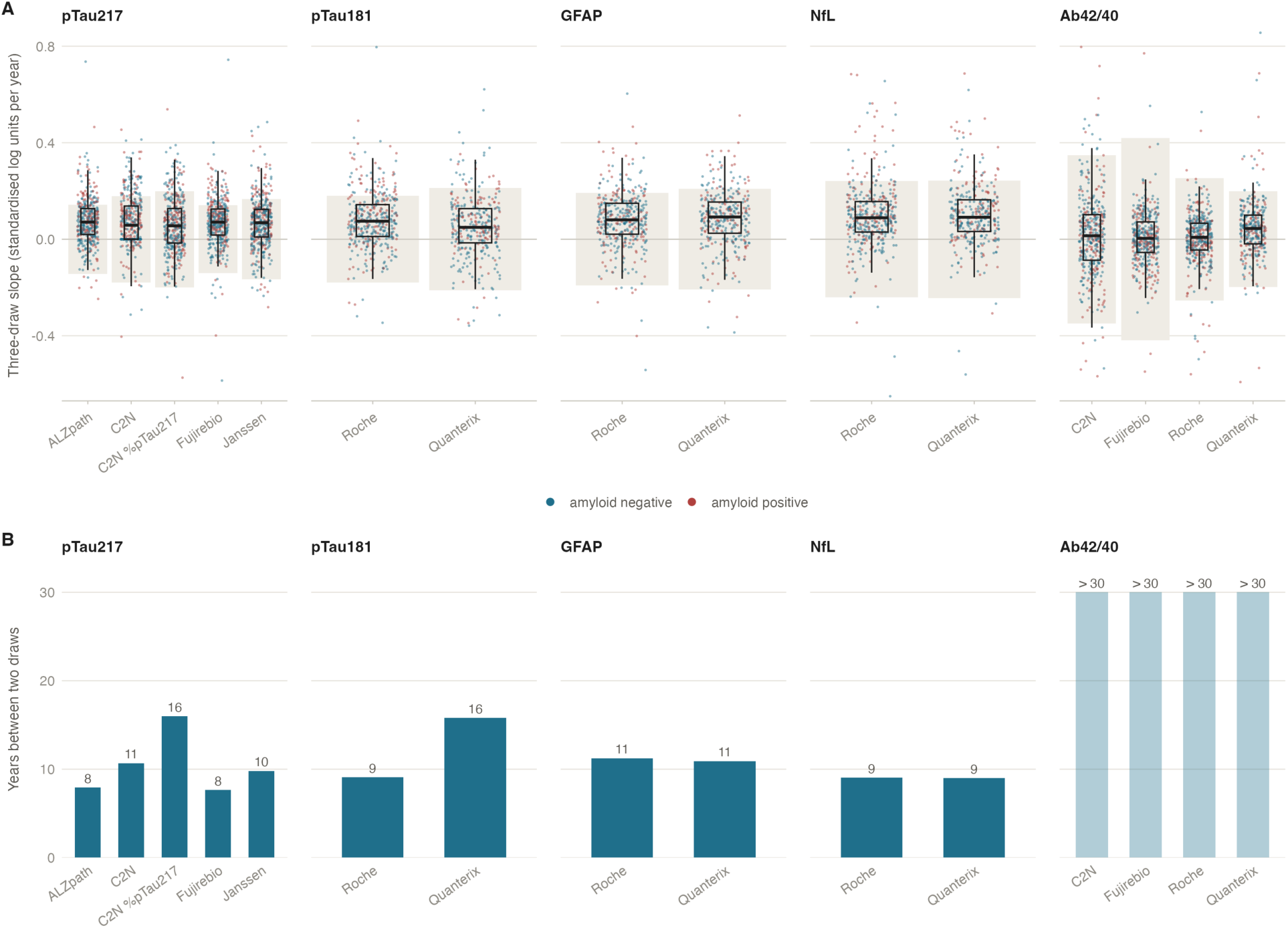
Individual change against measurement noise. (A) Per-participant three-sample slopes by platform (standardised log units per year), coloured by amyloid PET status at the first sample, with boxes showing the median and interquartile range. The shaded band is the 95% minimal detectable change for a three-sample slope on that platform, the range of slopes expected in 95% of people whose true rate of change is zero; slopes inside the band cannot be distinguished from no change. (B) Years between two plasma samples that a participant changing at the mean rate observed in amyloid-positive participants would need before the difference between the two measurements exceeds the 95% minimal detectable change for that platform; pale bars exceed 30 years.

**Table 3.** Minimal detectable change per platform on the delivered scale.

| Biomarker | Platform | Units | Median level | Within-person CV, % | Mean change per year, %: all | Mean change per year, %: amyloid positive | MDC95 between two measurements, % rise | MDC95, % fall | MDC95 of a three-sample slope, % per year | Years to exceed MDC95 | Slope above MDC95, % | Slope below −MDC95, % |
|---|---|---|---|---|---|---|---|---|---|---|---|---|
| pTau217 | ALZpath | pg/mL | 0.351 | 18 | 5.6 | 6.6 | 65 (54 to 79) | 40 (35 to 44) | 10 (8 to 12) | 8 (6 to 10) | 20 (14 to 28) | 2 |
| pTau217 | C2N | pg/mL | 1.76 | 31 | 7.1 | 8.1 | 131 (113 to 150) | 57 (53 to 60) | 18 (15 to 21) | 11 (9 to 14) | 17 (13 to 22) | 2 |
| pTau217 | C2N %pTau217 | % | 3.33 | 30 | 5.2 | 5.2 | 126 (108 to 144) | 56 (52 to 59) | 17 (15 to 20) | 16 (12 to 24) | 11 (8 to 18) | 1 |
| pTau217 | Fujirebio | pg/mL | 0.134 | 24 | 6.7 | 9.0 | 93 (77 to 112) | 48 (44 to 53) | 14 (12 to 16) | 8 (6 to 10) | 20 (13 to 27) | 3 |
| pTau217 | Janssen | pg/mL | 0.052 | 21 | 4.8 | 5.9 | 76 (64 to 88) | 43 (39 to 47) | 12 (10 to 14) | 10 (8 to 13) | 15 (11 to 21) | 2 |
| pTau181 | Roche | pg/mL | 1.05 | 15 | 3.6 | 4.6 | 51 (45 to 58) | 34 (31 to 37) | 8 (7 to 10) | 9 (7 to 12) | 17 (12 to 23) | 1 |
| pTau181 | Quanterix | pg/mL | 19 | 21 | 2.8 | 3.7 | 78 (65 to 93) | 44 (40 to 48) | 12 (10 to 14) | 16 (12 to 23) | 9 (6 to 13) | 3 |
| GFAP | Roche | pg/mL | 104 | 18 | 4.4 | 4.5 | 63 (53 to 74) | 39 (35 to 42) | 10 (8 to 12) | 11 (9 to 15) | 13 (9 to 19) | 2 |
| GFAP | Quanterix | pg/mL | 148 | 20 | 4.9 | 5.3 | 75 (64 to 84) | 43 (39 to 46) | 12 (10 to 13) | 11 (9 to 15) | 13 (8 to 18) | 3 |
| NfL | Roche | pg/mL | 3.68 | 19 | 4.0 | 6.0 | 69 (51 to 88) | 41 (34 to 47) | 11 (8 to 14) | 9 (7 to 11) | 10 (7 to 17) | 1 |
| NfL | Quanterix | pg/mL | 20 | 21 | 4.8 | 6.7 | 80 (61 to 97) | 44 (38 to 49) | 12 (10 to 15) | 9 (7 to 11) | 11 (8 to 18) | 1 |
| Aβ42/40 | C2N | ratio | 0.092 | 8 | −0.1 | 0.2 | 24 (20 to 27) | 19 (17 to 22) | 4 (4 to 5) |  | 4 (2 to 6) | 3 |
| Aβ42/40 | Fujirebio | ratio | 0.088 | 16 | 0.2 | 0.0 | 54 (26 to 90) | 35 (20 to 47) | 9 (4 to 13) |  | 1 (0 to 2) | 2 |
| Aβ42/40 | Roche | ratio | 0.122 | 8 | −0.1 | −0.1 | 25 (15 to 36) | 20 (13 to 26) | 5 (3 to 6) |  | 1 (1 to 4) | 3 |
| Aβ42/40 | Quanterix | ratio | 0.059 | 8 | −1.0 | −0.7 | 24 (19 to 31) | 20 (16 to 24) | 4 (3 to 6) | 32 (17 to 213) | 5 (3 to 9) | 2 |

Within-person CV is the coefficient of variation of a single measurement after removing the participant's linear trajectory, comprising analytical, pre-analytical and short-term biological variation. Mean change per year is from per-participant three-sample slopes, in all participants and in those amyloid positive at the first sample; negative values are decreases in the delivered direction (the Aβ42/40 ratio falls with pathology). MDC95 between two measurements is 1.96 × √2 × the within-person SD on the log scale, expressed as a percentage rise and as the equivalent percentage fall (one minus the reciprocal of the fold); MDC95 of a slope is 1.96 × the standard error of a three-sample slope at the median spread of sampling times. Years to exceed MDC95 is the number of years of compounding at the mean amyloid-positive rate of change needed to reach the two-point MDC95. The last two columns are the percentage of participants whose own three-sample slope lies above the slope MDC95 (a reliable increase) or below its negative (a reliable decrease). CV, coefficient of variation; MDC95, 95% minimal detectable change. The 95% intervals on the minimal detectable change, on the years to exceed it and on the percentage of participants above it are percentile intervals from 1,000 bootstrap resamples of participants. For Aβ42/40 the mean rate of change in amyloid-positive participants is not distinguishable from zero on three of the four platforms, so the interval between samples needed to exceed the minimal detectable change is not estimable and is left blank for those three platforms.

To determine how much within-person variation arose from the assays themselves, analytical CVs from quality-control samples were compared with within-person CVs (Supplementary Table S15). Quality-control CVs were 2–3% for Roche GFAP and NfL, 5–9% for other protein assays, and 11.5% for Fujirebio pTau217 (34% for its low control near the lower assay limit), versus within-person CVs of 15–31%. Analytical variation therefore accounted for 2% of within-person variance for Roche GFAP and NfL, 5–14% for pTau181 and Quanterix GFAP/NfL, and 3–23% for pTau217; the remaining non-analytical component corresponded to CVs of 14–30% across protein measures. Aβ42/40 was the exception: its 8% within-person CV on C2N, Roche, and Quanterix was largely explained by analytical CVs of 6–8% (97%, 90%, and 61% of variance, respectively).

To determine how large an individual change must be to exceed this variation, the minimal detectable change (MDC95) between two measurements was calculated. For pTau217 it ranged from 65% on ALZpath, 76% on Janssen, and 93% on Fujirebio to 131% on C2N pTau217; corresponding values were 51% and 78% for Roche and Quanterix pTau181, 63–80% for GFAP and NfL, and 24–54% for Aβ42/40. Paired differences showed no detectable difference between ALZpath and Janssen (10.40, 95% CI −2.68 to 21.48); ALZpath was lower than Fujirebio (27.86, 10.05–48.06), which was lower than C2N pTau217 (37.27, 13.16–60.71). Roche was lower than Quanterix for pTau181 (27.15, 14.90–40.80), GFAP (11.94, 1.10–19.90), and NfL (11.05, 0.57–19.21).

For pTau217, average amyloid-positive change took 8 years to exceed MDC95 on ALZpath or Fujirebio, 10 on Janssen, 11 on C2N pTau217, and 16 on C2N %pTau217. It took 9 years for Roche pTau181 and NfL on either platform, and 11 years for GFAP. For Aβ42/40, the time to exceed MDC95 was not estimable on three platforms because mean amyloid-positive change was indistinguishable from zero; on Quanterix it was 32 years (95% CI 17–213). Consistently, only 11–20% of participants had a three-sample pTau217 slope exceeding slope MDC95 (95% CIs 8–28%), compared with 9–17% for pTau181, GFAP, and NfL and 1–5% for Aβ42/40; only 1–3% showed a detectable decrease.

### Longer sampling spans improve reliability and reduce trial requirements

To determine how sampling schedule affects individual slope reliability, schedules were compared through the spread of sampling times (SST), the only design quantity entering the reliability equation (Figure 4A, Supplementary Table S16a). A third sample midway between two others added no reliability: three samples over four years equalled two (0.30 for single-platform pTau217), while front-and back-loaded schedules over the same span were equivalent (0.33). Two replicates at each end of four years (0.46) outperformed five annual samples (0.34), and two samples six years apart (0.49) outperformed every four-year schedule except six samples with three at each end (0.56). For one pTau217 platform, reliabilities of 0.70, 0.80, and 0.90 required spans of 9.4 (95% CI 8.4–10.8), 12.4 (11.0–14.1), and 18.5 (16.4–21.1) years with two or three samples; averaging five measures reduced these to 4.2 (3.7–4.8), 5.5 (4.9–6.3) and 8.3 (7.4–9.5) years if the variation shared across platforms at a draw is removed by averaging, and to 9.1 (7.5–11.3), 11.9 (9.8–14.8) and 17.8 (14.7– 22.2) years if it is not (Supplementary Table S16b). For reliability 0.70, corresponding spans were 7.5 (5.8–9.5) years for NfL, 8.2 (6.4–10.1) for GFAP, 9.1 (7.5–11.1) for pTau181, and 22.9 (15.3–42.3) for Aβ42/40.

**Figure 4.**
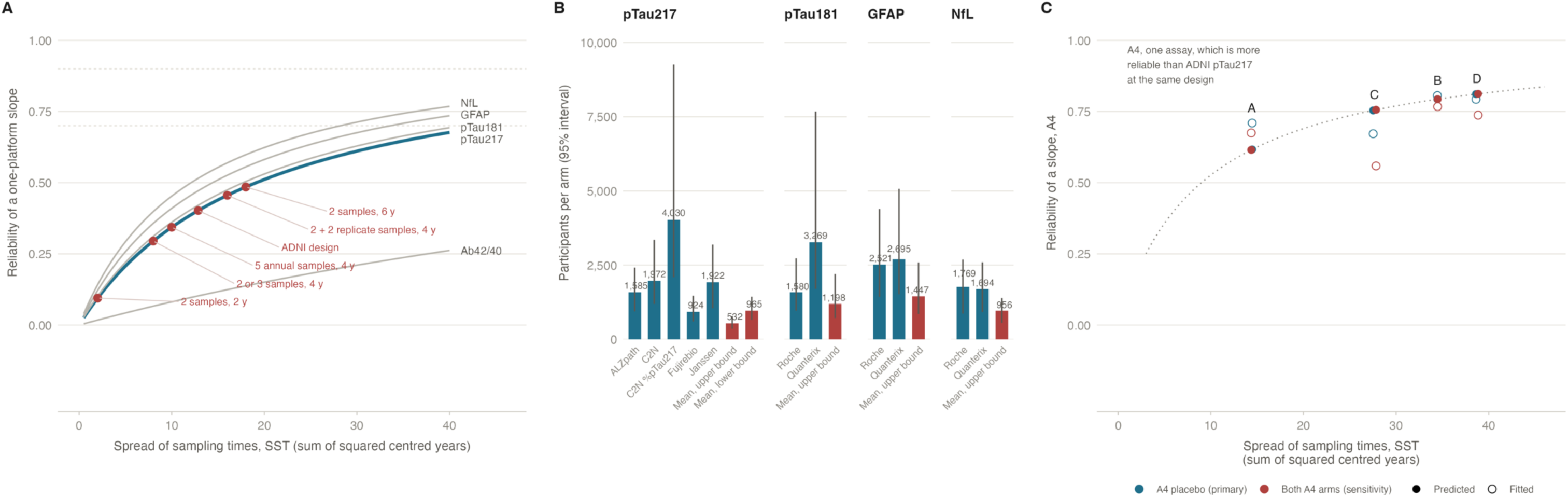
Consequences for sampling design and for trials. (A) Reliability of a one-platform slope as a function of the spread of sampling times (SST, the sum of squared deviations of a participant’s sampling times from their mean), which is the only feature of a schedule that reliability depends on. Curves are the five biomarkers (pTau217 in blue); points mark selected schedules for pTau217, including the observed ADNI design. Dotted lines mark reliabilities of 0.70 and 0.90. (B) Participants per arm required by a two-arm trial using the plasma slope as its outcome to detect a 25% slowing of the mean amyloid-positive rate of change over 24 months with three samples (two-sided α 0.05, 80% power), for each platform and for the mean of all platforms measuring that biomarker (orange) under the upper bound, in which averaging removes all occasion-level variation, and, for pTau217, the lower bound, in which the part shared across platforms at a blood draw remains. Vertical lines are 95% percentile intervals from 1,000 bootstrap resamples of participants. The calculation uses the amyloid-positive ADNI variance components and describes an ADNI-like population; Aβ42/40 is omitted because no platform gives a usable slope outcome. (C) The same axes tested within participants. In the A4 subgroup who gave a fourth plasma sample, each participant’s rate of change was recomputed from four subsets of their own samples: A the standard three samples over 4.8 years, B the same three samples over 7.3 years, C the same interval with the middle sample placed later, D all four samples. Filled points are the reliability the spread of sampling times predicts, open points the reliability refitted within these participants. The dotted line is the predicted relation for the A4 assay, which lies above the ADNI pTau217 curve in panel A because that assay is more reliable at the same design; the axes are otherwise identical to panel A.

To quantify the implications for trials using plasma slope as an outcome, sample sizes were calculated for 25% slowing of the amyloid-positive rate over 24 months with three samples, with the variance components estimated in amyloid-positive participants. For pTau217, participants required per arm were 924 (95% CI 598–1,468) for Fujirebio, 1,585 (940–2,414) for ALZpath, 1,922 (1,048– 3,194) for Janssen, 1,972 (1,193–3,351) for C2N pTau217, and 4,030 (2,091–9,256) for C2N %pTau217, falling to 532 (357–778) when all five measures were averaged if the variation shared at a draw is removed by averaging and to 965 (655–1,431) if it is not (Figure 4B, Supplementary Table S17). Under the upper bound, the pooled estimate required fewer participants than the best single measure (paired difference −392, 95% CI −806 to −138; under the lower bound 41, −353 to 381), whereas C2N %pTau217 required more than Fujirebio (ratio 4.36, 2.22 to 9.25), as did ALZpath (661 more, 23 to 1,285; Supplementary Table S17b). An 18-month four-sample schedule required 1,384 (891–2,205) to 6,241 (3,235–14,366) per arm (734 [491–1,076] to 1,509 [1,022–2,260] averaged), while five samples over 24 months reduced the requirements for single pTau217 measures by 17% (95% CI 15%–18%) for Fujirebio to 18% (18%–19%) for C2N %pTau217 compared with three; 50% slowing reduced all requirements four-fold. With change from baseline at the final sample as the endpoint instead of the slope, on whole-cohort variance components, the requirements were the same for this schedule and larger for the others. Roche pTau181 required 0.48 times as many participants as Quanterix (95% CI 0.25–0.88; 1,580 [961–2,730] vs 3,269 [1,699–7,672]), whereas NfL (1,769 [864–2,690] vs 1,694 [920–2,595]; paired difference 75, −502 to 632) and GFAP (2,521 [1,428–4,392] vs 2,695 [1,517–5,073]; −174, −1,434 to 872) showed no detectable difference between platforms. Aβ42/40 was not usable as a slope outcome.

### Reliable plasma change tracks different aspects of disease progression across biomarkers

To assess whether the reliable component of plasma change tracked disease progression, pooled slopes were correlated with concurrent change in the reference measures (Supplementary Figure S6, Table S18). Pooled pTau217 correlated with amyloid PET change (Spearman ρ=0.39, 95% CI 0.30– 0.48; single platforms 0.23–0.36, Supplementary Table S19) and CSF pTau181/Aβ42 change (0.35, 0.21–0.47), but weakly or not with cognition (0.05 [−0.06 to 0.15] to 0.12 [0.01–0.22]) or hippocampal volume (0.11, −0.00 to 0.22). NfL showed the opposite pattern, correlating with ADAS-Cog13 (0.22, 0.12–0.33), mPACC (0.19, 0.08–0.30), MMSE (0.14, 0.04–0.25), and hippocampal volume (0.20, 0.09–0.31), but not detectably with amyloid PET (0.05, −0.06 to 0.14). pTau181 and GFAP correlated with amyloid PET (0.28 [0.18–0.36] and 0.21 [0.11–0.31]) and CSF change (0.24 [0.09–0.38] and 0.24 [0.10–0.37]), but weakly or not with cognition. Aβ42/40 showed no detectable association with any reference measure except ADAS-Cog13 (−0.16, 95% CI −0.32 to −0.01), in the direction opposite to worsening. The pooled pTau217 slope correlated with amyloid PET change at 0.39, against 0.36 for the best single platform (ALZpath; difference 0.03, 95% CI −0.03 to 0.09) and 0.33 for Janssen (difference 0.06, −0.00 to 0.13), and exceeded C2N pTau217, C2N %pTau217 and Fujirebio by 0.08 (0.01–0.15) to 0.16 (0.08–0.24); pooled slopes were not more strongly related than the best single platform for every biomarker and reference measure; no platform drove the pooled pTau217 estimate, as omitting any one left slopes correlated ≥0.97 with the full pooled slope; for Aβ42/40 these correlations were 0.87–0.95 (Supplementary Table S20). For pTau217, validity against Centiloid change was 0.44 (0.32–0.54) in amyloid-negative and 0.21 (0.05–0.37) in amyloid-positive participants (difference −0.22, 95% CI −0.42 to −0.03).

### Aβ42/40 change is insufficiently reliable to distinguish stability from measurement noise

To determine whether apparent longitudinal stability of Aβ42/40 reflected biology or limited measurement, its reproducibility was examined across analyses. Aβ42/40 was consistently the outlier: generalisability was 0.32 for one measurement, slope reliability 0.10 on one platform and 0.31 with four platforms averaged, and change showed no detectable association with the reference measures except ADAS-Cog13. Although its absolute cross-platform limits of agreement were narrow (SD of log differences 0.17–0.23 vs 0.15 for NfL), its intraclass correlations were lowest because its between-participant CV is only 10–17%. Thus, measurement error is comparable to the biological range, so apparent stability cannot be distinguished from inability to measure individual change.

### A4/LEARN replicates slope reliability and confirms the importance of sampling span

To test whether individual slope reliability replicated in an independent cohort and assay, pTau217 was analysed in 1,093 A4/LEARN participants measured on one platform over up to four samples.

Reliability was 0.54 (95% CI 0.46–0.61), versus 0.40 (0.34–0.46) for single-platform pTau217 in ADNI, and 0.30 (0.16–0.42) to 0.55 (0.33–0.67) for the ADNI pTau217 immunoassays fitted alone, despite similar spread of sampling times (SST 13.4 vs 12.9 years²). Reliability was 0.60 (0.50–0.68) in A4 placebo participants, all with elevated amyloid at screening, and 0.35 (0.21–0.48) in LEARN, reproducing the ADNI finding that reliability is higher where there is more true change to detect. Mean amyloid-positive change was 7.0%/year, within the 5.2–9.0% ADNI range. MDC95 was 46% (Supplementary Table S21), requiring 5.6 years for average amyloid-positive change to exceed it. Three-sample increases exceeded slope MDC95 in 29% overall and 39% of A4 placebo participants, versus 11–20% in ADNI, consistent with the greater proportion of amyloid-positive participants.

To test whether variation around the fitted trajectory reflected measurement variation rather than curvature, 1,020 baseline/week-12 pairs (median 85 days) provided a direct short-interval estimate. Within-person CV was 14.8% (95% CI 13.7–16.0), ICC(2,1) 0.90 (0.88–0.91), and limits of agreement −32% to 53% (Supplementary Figure S7); 5.0% differed by more than the 50% MDC95 despite little expected biological change over 12 weeks. The three-sample model residual variance was 1.02 times the 12-week within-person variance (Supplementary Table S22), as expected by construction because the week-12 sample is one of the three fitted; the test therefore used the participants with a fourth sample. In the 146 participants with one, the residual around a line through the baseline, week-240 and extension samples was 1.57 times the 12-week variance (95% CI 1.06–2.42), so 12-week variation accounted for 64% (41–95%) of the variation around a seven-year line; in the 77 placebo participants the ratio was 1.44 (0.91–2.40), with an interval that included 1 (Supplementary Table S23a). There was no detectable curvature of the mean trajectory: the quadratic term was −0.0015 per year² (95% CI −0.0061 to 0.0031), which excludes a departure from a straight line larger than 0.08 standardised units at the ends of the 7.3-year window.

To test the predicted effect of sampling design for the same participants and assay, slopes were recomputed from subsets of each participant’s samples (Supplementary Table S23b). Slopes from only the baseline and week-12 samples had a predicted reliability of 0.002. As predicted, these slopes correlated at only 0.02 with concurrent change in amyloid PET. The same participants’ slopes from all three samples, spread over several years, gave a correlation of 0.17. The two widely spaced pairs, with predicted reliabilities of 0.51 and 0.48, gave correlation ratios of 0.98 and 0.83 versus 0.95 and 0.92 predicted, and 1.05 and 0.82 with Pearson correlation (Supplementary Figure S8). Thus, with participant and assay fixed, validity changed with SST as predicted.

Between-participant comparisons showed a steeper gradient in ADNI, but no consistent gradient in A4/LEARN, where the bands of low SST held 3 to 57 participants (Supplementary Table S23c). In ADNI, the lowest SST fifth had a pTau217 slope-to-Centiloid correlation 0.34 times that of the highest fifth, versus 0.67 predicted from plasma reliability alone (Supplementary Table S24). Their Centiloid slopes were also less precise, covering 3.9 years with three scans versus 8.0 years with five; attenuation on both sides predicted a ratio of 0.45, explaining most of the discrepancy. Thus, between-participant comparisons confound plasma sampling design with reference-measure precision, making the within-participant analysis the stronger test.

Finally, to separate sampling span from sample number, four-sample A4 participants were compared within person (Figure 4C, Supplementary Table S23d). Among 77 placebo participants, extending three samples from 4.8 to 7.3 years increased SST from 14.5 to 34.5 years², predicted reliability from 0.62 to 0.79, fitted reliability from 0.71 to 0.81 (difference 0.10, 95% CI 0.03–0.22), and changed the correlation with Centiloid change by 0.05 (−0.12 to 0.21). Over the same 7.3-year interval, adding a fourth sample increased predicted reliability by only 0.02 and had no detectable effect on fitted reliability (−0.01, −0.08 to 0.05) or Centiloid correlation (0.00, −0.04 to 0.05); moving the middle sample from week 12 to week 240 (4.6 years) reduced SST and changed fitted reliability by −0.13 (−0.32 to 0.00) and the Centiloid correlation by 0.02 (−0.06 to 0.10). Model-free comparison agreed: the long-interval three-sample slope correlated 0.98 with the four-sample slope versus 0.86 for the standard three-sample slope. The same pattern of reliability occurred in all 146 participants from both arms.

## Discussion

Cross-sectional agreement about a person’s biomarker level at a single timepoint did not imply reliable measurement of that person’s change. Agreement about level was high for pTau217, pTau181, GFAP, and NfL, reproducing previous head-to-head comparisons [8], but agreement about individual rates of change was substantially lower. With three samples over a median 5.0 years, slope reliability was at most 0.40 for pTau217, 0.42–0.52 for the other three proteins, and 0.10 for Aβ42/40. The same measurement variation is small relative to between-person differences in level (generalisability coefficients 0.70–0.81 for the four proteins) but large relative to between-person differences in change: true rates differed by an SD of only 0.07–0.10 standardised units per year, against a measurement SD of 0.30–0.35 units at each sample.

This disagreement did not reflect systematic differences between platforms. Platform-specific slope variance was approximately zero for each biomarker, and an effect at the threshold detectable with 80% power would reduce pTau217 reliability only from 0.40 to 0.39; smaller effects cannot be excluded but would explain little of the disagreement. The main limitation is random variation at each sampling occasion, which recalibration cannot remove but additional independent measurements can reduce. For GFAP and NfL, the two platforms agreed closely about change (slope correlations 0.81 and 0.88), yet each assay fitted alone gave reliabilities of only 0.03 to 0.19 for GFAP and 0.01 to 0.10 for NfL. This is consistent with variation shared by both platforms at each blood draw; its size cannot be estimated with two platforms, and its biological or pre-analytical origin cannot be determined from this design.

In A4/LEARN, slope reliability was 0.54 on a sixth assay, so limited reliability was not specific to ADNI. In participants with a fourth sample, variance between samples 12 weeks apart accounted for 64% (95% CI 41–95%) of the residual variance around seven-year trajectories, with no detectable curvature in the mean trajectory. Because little true pTau217 change is expected over 12 weeks [16,18], much of the unexplained long-term variation appears to be short-interval measurement variation; the remainder could reflect slower biological variation, individual curvature, or response to open-label treatment. The 12-week ICC of 0.90 was consistent with the correlation of 0.91 reported for repeats 12 weeks to 1.75 years apart [18].

### Relation to previous work

Previous longitudinal studies established the biology and prognostic value of plasma change. In the same ADNI samples, pTau217 increases were associated with amyloid accumulation, with a similar ordering of assays by rate of change [6], and with cognitive decline, depending on disease stage and assay [7]. In the same cohort, baseline values were more strongly associated with subsequent change in amyloid PET, cortical thickness, and cognition than were the biomarkers’ own rates of change [44]. A single-assay series cannot show how much of one person’s observed slope is signal. By comparing several assays on aliquots of the same longitudinal samples, the present analysis separates shared change from platform-specific deviations and translates reliability into minimal detectable change, sampling requirements, and trial sizes.

### Reliable rates of change track different aspects of disease progression

Low reliability weakens, but does not preclude, associations with concurrent markers of disease progression. The pooled multi-platform slope of pTau217 was associated with change in amyloid PET and CSF pTau181/Aβ42, whereas that of NfL was associated with hippocampal atrophy and cognitive decline. In amyloid-stratified analyses, pTau217 slope was more strongly associated with amyloid accumulation in amyloid-negative than amyloid-positive participants, as also reported by Yakoub et al. [6], despite higher slope reliability in the positive group. This weaker association cannot be explained by poorer measurement and is consistent with a weaker link between pTau217 change and further amyloid accumulation after amyloid positivity.

### Implications for individual monitoring and trials

For pTau217, measurements had to change by 65–131%, depending on the measure, to exceed measurement noise. These thresholds are wider than reference change values from repeat sampling over weeks (−28% to 38% for ALZpath pTau217 in healthy adults sampled weekly [16]; from −15%/17% for Aβ42/40 to −30%/42% for pTau181 in memory-clinic patients sampled within 36 days [17]), in part because they include all variation around a five-year trajectory. At mean amyloid-positive rates, even the best measure needed eight years for average change to exceed this threshold, and only 11–20% of participants had a three-sample pTau217 increase exceeding the slope threshold.

Reliability depends on sampling design only through SST, which grows with the square of the span, so span has more effect than frequency: doubling the span quadruples SST, whereas increasing from two to six evenly spaced samples within a fixed span increases it only 1.40-fold. Placement also matters: two samples at each end of a span give 1.80 times the SST of four evenly spaced samples and 1.60 times that of five annual samples, so endpoint replication over four years gave a reliability of 0.46 against 0.34 for five annual samples. In A4, extending three samples from 4.8 to 7.3 years substantially increased reliability, whereas a fourth sample over the same span added little.

Averaging replicate measurements at each occasion also improves reliability without changing SST. With independent errors, averaging m replicates is equivalent to extending the span by √m (41% for two measurements, 124% for five). The gain is smaller when variation is shared across replicates, as with aliquots of one blood draw: for pTau217 the shared part was 30% (95% CI 25%–36%) of occasion-level variation. Because analytical imprecision was a minority of within-person variation for the protein biomarkers, longer spans, endpoint replication, and averaging offer greater gains than modest improvements in assay precision. These relationships assume linear trajectories and complete follow-up, and the longest spans extrapolate beyond the median 5.0 years observed in ADNI.

The same problem affects trials using plasma slope as an outcome. In an ADNI-like population, detecting 25% slowing of pTau217 change over 24 months required 924–4,030 participants per arm across measures, and 532–965 with averaged measurements. With 146 amyloid-positive participants and three samples each, the intervals are wide: a 4.36-fold difference between measures is resolved, but a 1.72-fold difference between two good assays (Fujirebio 924 against ALZpath 1,585) only just. Change from baseline to the final sample, on whole-cohort components, gave the same or larger requirements. Ferreira et al. [4] reported smaller requirements in amyloid-positive and enriched populations using single-assay pTau217. These estimates concern slowing of the natural rate of change, not treatment-induced reductions in level, which can appear much earlier [5].

### Aβ42/40 is an important exception

The Aβ42/40 generalisability coefficient was 0.32 and its single-platform slope reliability 0.10, rising only to 0.31 with four platforms averaged; the ratio varied by only 10–17% between people, so measurement variation was large relative to its biological range. Its change showed no detectable association with the reference measures except ADAS-Cog13, so its apparent longitudinal stability, here and previously [6], cannot be distinguished from limited ability to measure change. Unlike the protein biomarkers, most within-person Aβ42/40 variation on platforms with quality-control data was analytical, so better assay precision or technical replication could address this.

### Strengths and limitations

The main strength is the longitudinal multi-platform design, which separates variation shared across assays, including participant level and change, from platform-specific variation. Reliabilities were stable across treatments of quantification limits, covariate adjustment, analysis scales except untransformed NfL, and alternative models. Replication in 1,093 A4/LEARN participants and 12-week repeat measurements tested assumptions that ADNI alone could not. However, pre-analytical variation shared across platforms is counted as change in this design, so the reliabilities are upper bounds and may be optimistic for real-world monitoring. For pTau217, the averaged reliability lies between 0.42 (0.32–0.52), counting this variation as error, and 0.77 (0.72–0.81), counting it as change; for the other biomarkers only the upper bound is estimable, and repeat blood draws days apart on the same platforms are needed to separate it. With three ADNI samples per participant, shared occasion variation is not cleanly separable from the slope, so the gain from averaging is less certain than single-assay reliability.

The four-sample test rests on 146 A4 participants whose fourth sample was taken during open-label treatment in both arms, and its placebo-arm interval includes one. Storage time, 5.5 to 13.6 years, showed no detectable association with measured levels but is collinear with enrolment period and assay batch, and freeze–thaw cycles were not recorded; the implied bias in the rate of change could lie within −67% to 14% of the mean amyloid-positive rate for ALZpath pTau217. A steady degradation bias would shift mean rates and derived quantities, not reliabilities or minimal detectable change. Three platforms reported duplicate means and three single measurements, with similar within-person variation, and quality-control data separate analytical from other variation only approximately. C2N pTau217 was at its quantification limit in 36% of observations; sensitivity analyses did not materially alter joint-model reliabilities, and excluding these samples lowered its minimal detectable change from 131% to 101%. ADNI is a research cohort, and the cross-platform decomposition rests on ADNI alone; A4/LEARN replicated the reliability and sampling-design findings on one assay.

## Conclusion

Plasma assays agree substantially more about a person’s level than about their rate of change, and disagreement about change is predominantly random rather than systematic; short-term variation accounted for 64% (41%–95%) of the seven-year model residual. With three samples over a median 5.0 years, at most 0.40–0.52 of individual slope variation on one assay was reliable for the four protein biomarkers in ADNI, and 0.54 for pTau217 in A4/LEARN. Reliability improved with longer spans, endpoint replication, and averaging measures, and for pTau217 also depended on the assay, whose minimal detectable change ranged from 65% to 131% and trial sizes from 924 to 4,030 per arm. Aβ42/40 change had little useful reliability. These results quantify the detectable change and sampling required for individual serial testing and the sample sizes required for trials using plasma change as an outcome.

## Data Availability

Data collection and sharing for the Alzheimer′s Disease Neuroimaging Initiative (ADNI) is funded by the National Institute on Aging (National Institutes of Health Grant U19AG024904). The grantee organization is the Northern California Institute for Research and Education. In the past, ADNI has also received funding from the National Institute of Biomedical Imaging and Bioengineering, the Canadian Institutes of Health Research, and private sector contributions through the Foundation for the National Institutes of Health (FNIH) including generous contributions from the following: AbbVie, Alzheimer′s Association; Alzheimer′s Drug Discovery Foundation; Araclon Biotech; BioClinica, Inc.; Biogen; Bristol-Myers Squibb Company; CereSpir, Inc.; Cogstate; Eisai Inc.; Elan Pharmaceuticals, Inc.; Eli Lilly and Company; EuroImmun; F. Hoffmann-La Roche Ltd and its affiliated company Genentech, Inc.; Fujirebio; GE Healthcare; IXICO Ltd.; Janssen Alzheimer Immunotherapy Research & Development, LLC.; Johnson & Johnson Pharmaceutical Research & Development LLC.; Lumosity; Lundbeck; Merck & Co., Inc.; Meso Scale Diagnostics, LLC.; NeuroRx Research; Neurotrack Technologies; Novartis Pharmaceuticals Corporation; Pfizer Inc.; Piramal Imaging; Servier; Takeda Pharmaceutical Company; and Transition Therapeutics. The plasma measures analysed here were generated by the Foundation for the National Institutes of Health (FNIH) Biomarkers Consortium project Plasma Aβ and Phosphorylated Tau as Predictors of Amyloid and Tau Positivity in Alzheimer′s Disease. The authors thank the FNIH Biomarkers Consortium project team and the participating assay developers for making these measures available through ADNI. Data used in the preparation of this article were also obtained from the Anti-Amyloid Treatment in Asymptomatic Alzheimer′s disease (A4) study and its companion observational study (LEARN), obtained through the A4 study data repository. The A4 and LEARN investigators contributed data but did not participate in the analysis or writing of this report. The A4 Study was funded by a public-private-philanthropic partnership, including funding from the National Institutes of Health-National Institute on Aging (R01AG063689; U19AG010483; U24AG057437), Eli Lilly and Company, Alzheimer′s Association, Accelerating Medicines Partnership through the Foundation for the National Institutes of Health, GHR Foundation, Davis Alzheimer Prevention Program, an anonymous foundation and additional private donors to the Brigham and Women′s Hospital, with in-kind support from Avid, Cogstate, Albert Einstein College of Medicine, and the Foundation for Neurologic Diseases. The companion observational Longitudinal Evaluation of Amyloid Risk and Neurodegeneration (LEARN) Study was funded by the Alzheimer′s Association and GHR Foundation. We thank the A4 and LEARN Study Teams, current and former, and all study participants.

Both datasets are available to qualified investigators on application to their respective repositories; the authors are not permitted to redistribute them. Analysis code reproducing every table and figure in this report is available at https://github.com/oxford1869/ad-plasma-biomarker-rates-of-change.

https://adni.loni.usc.edu/data-samples/adni-data/

https://www.a4studydata.org/

## Acknowledgements

funding and data availability

## Author contributions

YY conceived and designed the study, curated and analysed the data, and wrote the first draft. IK and VR supervised the work. JR, KKP, AB, RB and RM-S reviewed and edited the manuscript. All authors read and approved the final manuscript.

## Competing interests

IK has received grant funding for an investigator-initiated study from Novo Nordisk. He has received consultation fees from Novo Nordisk, Zylorion, Johnson and Johnson. IK has received speaker fees for non-promotional educational events by Novo Nordisk and Eisai. IK is a paid medical advisor for medical device companies in the dementia field (Five Lives, Prima Mente, Oxford Brain Diagnostics). All other authors declare no competing interests.

Data collection and sharing for the Alzheimer’s Disease Neuroimaging Initiative (ADNI) is funded by the National Institute on Aging (National Institutes of Health Grant U19AG024904). The grantee organization is the Northern California Institute for Research and Education. In the past, ADNI has also received funding from the National Institute of Biomedical Imaging and Bioengineering, the Canadian Institutes of Health Research, and private sector contributions through the Foundation for the National Institutes of Health (FNIH) including generous contributions from the following: AbbVie, Alzheimer’s Association; Alzheimer’s Drug Discovery Foundation; Araclon Biotech; BioClinica, Inc.; Biogen; Bristol-Myers Squibb Company; CereSpir, Inc.; Cogstate; Eisai Inc.; Elan Pharmaceuticals, Inc.; Eli Lilly and Company; EuroImmun; F. Hoffmann-La Roche Ltd and its affiliated company Genentech, Inc.; Fujirebio; GE Healthcare; IXICO Ltd.; Janssen Alzheimer Immunotherapy Research & Development, LLC.; Johnson & Johnson Pharmaceutical Research & Development LLC.; Lumosity; Lundbeck; Merck & Co., Inc.; Meso Scale Diagnostics, LLC.; NeuroRx Research; Neurotrack Technologies; Novartis Pharmaceuticals Corporation; Pfizer Inc.; Piramal Imaging; Servier; Takeda Pharmaceutical Company; and Transition Therapeutics. The plasma measures analysed here were generated by the Foundation for the National Institutes of Health (FNIH) Biomarkers Consortium project Plasma Aβ and Phosphorylated Tau as Predictors of Amyloid and Tau Positivity in Alzheimer’s Disease. The authors thank the FNIH Biomarkers Consortium project team and the participating assay developers for making these measures available through ADNI.

Data used in the preparation of this article were also obtained from the Anti-Amyloid Treatment in Asymptomatic Alzheimer’s disease (A4) study and its companion observational study (LEARN), obtained through the A4 study data repository. The A4 and LEARN investigators contributed data but did not participate in the analysis or writing of this report. The A4 Study was funded by a public-private-philanthropic partnership, including funding from the National Institutes of Health-National Institute on Aging (R01AG063689; U19AG010483; U24AG057437), Eli Lilly and Company, Alzheimer’s Association, Accelerating Medicines Partnership through the Foundation for the National Institutes of Health, GHR Foundation, Davis Alzheimer Prevention Program, an anonymous foundation and additional private donors to the Brigham and Women’s Hospital, with in-kind support from Avid, Cogstate, Albert Einstein College of Medicine, and the Foundation for Neurologic Diseases. The companion observational Longitudinal Evaluation of Amyloid Risk and Neurodegeneration (LEARN) Study was funded by the Alzheimer’s Association and GHR Foundation. We thank the A4 and LEARN Study Teams, current and former, and all study participants.

AI tools were used to assist with language editing, improving clarity, and cross-checking reported numerical values against the original statistical analysis output files. All statistical analyses were specified, conducted, and verified by the authors. AI tools were not used to generate or modify research data or analytical results, nor did they influence the study’s scientific interpretations or conclusions. The authors reviewed and verified the final manuscript and take full responsibility for its content.

## Ethics

This is a secondary analysis of de-identified data from two completed studies, each of which obtained its own ethical approval and written informed consent. ADNI was approved by the institutional review board of every participating site, and all participants, or their study partner where appropriate, gave written informed consent at enrolment. The A4 study and its companion observational study LEARN were likewise approved at every participating site, and all participants gave written informed consent. No new data were collected for the present analysis and no participant was contacted. Analysis of these de-identified datasets was exempt from further ethical review, and access was granted under the data use agreements of the two repositories.

## Reporting guideline

This report follows the Guidelines for Reporting Reliability and Agreement Studies (GRRAS) [45]; the completed checklist is Supplementary Table S25.

## References

1. Palmqvist S, Whitson HE, Allen LA, et al. Alzheimer’s Association clinical practice guideline on the use of blood-based biomarkers in the diagnostic workup of suspected Alzheimer’s disease within specialized care settings. Alzheimers Dement. 2025;21(7):e70535.

2. Schöll M, Verberk IMW, del Campo M, et al. Challenges in the practical implementation of blood biomarkers for Alzheimer’s disease. Lancet Healthy Longev. 2024;5(10):100630.

3. Petersen KK, et al. Predicting onset of symptomatic Alzheimer’s disease with plasma p-tau217 clocks. Nat Med. 2026;32(3):1085–1094.

4. Ferreira PCL, Bellaver B, Povala G, et al. Evaluating plasma p-tau217 as an endpoint for Alzheimer disease clinical trials. Neurology. 2026;106(1):e214441. doi:10.1212/WNL.0000000000214441.

5. Pontecorvo MJ, Lu M, Burnham SC, Schade AE, Dage JL, Shcherbinin S, et al. Association of donanemab treatment with exploratory plasma biomarkers in early symptomatic Alzheimer disease: a secondary analysis of the TRAILBLAZER-ALZ randomized clinical trial. JAMA Neurol. 2022;79(12):1250–1259. doi:10.1001/jamaneurol.2022.3392.

6. Yakoub Y, Qiu T, Peyrot C, Salvadó G, Villeneuve S, Pichet Binette A. Trajectories of plasma biomarkers, amyloid-beta burden and cognitive decline in Alzheimer’s disease: a longitudinal ADNI study. medRxiv. 2025; doi:10.1101/2025.09.30.25337003. PMID 41256144.

7. Ahn J, Yun J, Weiner MW, et al. Longitudinal plasma p-tau217 as a marker for tracking progression and predicting cognitive decline in Alzheimer’s disease. Alzheimers Res Ther. 2026;18:170.

8. Schindler SE, Petersen KK, Saef B, et al. Head-to-head comparison of leading blood tests for Alzheimer’s disease pathology. Alzheimers Dement. 2024;20(11):8074–8096.

9. Ashton NJ, Keshavan A, Brum WS, et al. The Alzheimer’s Association Global Biomarker Standardization Consortium (GBSC) plasma phospho-tau Round Robin study. Alzheimers Dement. 2025;21(2):e14508.

10. Nunnally JC, Bernstein IH. Psychometric Theory. 3rd ed. New York: McGraw-Hill; 1994.

11. Spearman C. The proof and measurement of association between two things. Am J Psychol. 1904;15(1):72–101.

12. Rogosa D, Brandt D, Zimowski M. A growth curve approach to the measurement of change. Psychol Bull. 1982;92(3):726–748.

13. Willett JB. Some results on reliability for the longitudinal measurement of change: implications for the design of studies of individual growth. Educ Psychol Meas. 1989;49(3):587–602.

14. Brennan RL. Generalizability Theory. New York: Springer; 2001.

15. Campbell DT, Fiske DW. Convergent and discriminant validation by the multitrait-multimethod matrix. Psychol Bull. 1959;56(2):81–105.

16. Brum WS, Ashton NJ, Simrén J, et al. Biological variation estimates of Alzheimer’s disease plasma biomarkers in healthy individuals. Alzheimers Dement. 2024;20(2):1284–1297. doi:10.1002/alz.13518.

17. Clemmensen FK, Gramkow MH, Simonsen AH, et al. Short-term variability of Alzheimer’s disease plasma biomarkers in a mixed memory clinic cohort. Alzheimers Res Ther. 2025;17(1):26. doi:10.1186/s13195-024-01658-7.

18. Rissman RA, Donohue MC, Langford O, et al. Longitudinal phospho-tau217 predicts amyloid positron emission tomography in asymptomatic Alzheimer’s disease. J Prev Alzheimers Dis. 2024;11(4):823–830. doi:10.14283/jpad.2024.134.

19. Sperling RA, Donohue MC, Raman R, et al. Trial of solanezumab in preclinical Alzheimer’s disease. N Engl J Med. 2023;389(12):1096–1107. doi:10.1056/NEJMoa2305032.

20. Navitsky M, Joshi AD, Kennedy I, et al. Standardization of amyloid quantitation with florbetapir standardized uptake value ratios to the Centiloid scale. Alzheimers Dement. 2018;14(12):1565–1571. doi:10.1016/j.jalz.2018.06.1353.

21. Shaw LM, Korecka M, Lee EB, Cousins KAQ, Vanderstichele H, Schindler SE, et al. ADNI Biomarker Core: a review of progress since 2004 and future challenges. Alzheimers Dement. 2025;21(1):e14264. doi:10.1002/alz.14264.

22. Klunk WE, Koeppe RA, Price JC, et al. The Centiloid Project: standardizing quantitative amyloid plaque estimation by PET. Alzheimers Dement. 2015;11(1):1–15.e4.

23. Jack CR Jr, Bernstein MA, Fox NC, et al. The Alzheimer’s Disease Neuroimaging Initiative (ADNI): MRI methods. J Magn Reson Imaging. 2008;27(4):685–691. doi:10.1002/jmri.21049.

24. Rosen WG, Mohs RC, Davis KL. A new rating scale for Alzheimer’s disease. Am J Psychiatry. 1984;141(11):1356–1364. doi:10.1176/ajp.141.11.1356.

25. Mohs RC, Knopman D, Petersen RC, et al. Development of cognitive instruments for use in clinical trials of antidementia drugs: additions to the Alzheimer’s Disease Assessment Scale that broaden its scope. Alzheimer Dis Assoc Disord. 1997;11(Suppl 2):S13–S21. doi:10.1097/00002093-199700112-00003.

26. Donohue MC, Sperling RA, Salmon DP, et al. The preclinical Alzheimer cognitive composite: measuring amyloid-related decline. JAMA Neurol. 2014;71(8):961–970.

27. Folstein MF, Folstein SE, McHugh PR. “Mini-mental state”: a practical method for grading the cognitive state of patients for the clinician. J Psychiatr Res. 1975;12(3):189–198.

28. Royse SK, Minhas DS, Lopresti BJ, et al. Validation of amyloid PET positivity thresholds in centiloids: a multisite PET study approach. Alzheimers Res Ther. 2021;13:99. doi:10.1186/s13195-021-00836-1.

29. R Core Team. R: A Language and Environment for Statistical Computing. Vienna: R Foundation for Statistical Computing; 2025.

30. Bates D, Mächler M, Bolker B, Walker S. Fitting linear mixed-effects models using lme4. J Stat Softw. 2015;67(1):1–48.

31. Proust-Lima C, Philipps V, Liquet B. Estimation of extended mixed models using latent classes and latent processes: the R package lcmm. J Stat Softw. 2017;78(2):1–56.

32. Bland JM, Altman DG. Statistical methods for assessing agreement between two methods of clinical measurement. Lancet. 1986;1(8476):307–310.

33. Shrout PE, Fleiss JL. Intraclass correlations: uses in assessing rater reliability. Psychol Bull. 1979;86(2):420–428.

34. Cohen J. A coefficient of agreement for nominal scales. Educ Psychol Meas. 1960;20(1):37–46. doi:10.1177/001316446002000104.

35. Laird NM, Ware JH. Random-effects models for longitudinal data. Biometrics. 1982;38(4):963–974.

36. Self SG, Liang KY. Asymptotic properties of maximum likelihood estimators and likelihood ratio tests under nonstandard conditions. J Am Stat Assoc. 1987;82(398):605–610. doi:10.1080/01621459.1987.10478472.

37. Stram DO, Lee JW. Variance components testing in the longitudinal mixed effects model. Biometrics. 1994;50(4):1171–1177.

38. Efron B, Tibshirani RJ. An Introduction to the Bootstrap. New York: Chapman and Hall; 1993.

39. Bland JM, Altman DG. Statistics notes: measurement error. BMJ. 1996;313(7059):744.

40. Koo TK, Li MY. A guideline of selecting and reporting intraclass correlation coefficients for reliability research. J Chiropr Med. 2016;15(2):155–163.

41. Rubin DB. Multiple Imputation for Nonresponse in Surveys. New York: Wiley; 1987.

42. Weir JP. Quantifying test-retest reliability using the intraclass correlation coefficient and the SEM. J Strength Cond Res. 2005;19(1):231–240.

43. Raudenbush SW, Liu XF. Effects of study duration, frequency of observation, and sample size on power in studies of group differences in polynomial change. Psychol Methods. 2001;6(4):387–401. doi:10.1037/1082-989X.6.4.387.

44. Saef B, Petersen KK, Volluz K, et al. Exploring longitudinal relationships among Alzheimer’s disease biomarkers. Alzheimers Dement. 2026;22(8):e71711. doi:10.1002/alz.71711.

45. Kottner J, Audigé L, Brorson S, et al. Guidelines for Reporting Reliability and Agreement Studies (GRRAS) were proposed. J Clin Epidemiol. 2011;64(1):96–106. doi:10.1016/j.jclinepi.2010.03.002.

